# Hearing Loss as a Modifiable Risk Factor for Dementia

**DOI:** 10.64898/2026.03.16.26348518

**Authors:** Abigail Dichter, Ella J. Lee, Timothy Park, Karen Tawk, Elham Ghanbarian, Mehdi Abouzari

## Abstract

This study evaluated hearing loss (HL) as a potential early indicator of dementia. We analyzed 16,270 participants from the *All of Us* database (1980 to 2022), including 1,224 (7.5%) with dementia, matched to controls by U.S. census demographics. Severe survey-reported HL showed the strongest association with dementia (odds ratio [OR] 6.76), followed by sensorineural HL (SNHL) (OR 3.90), smoking (OR 1.71), parental HL (OR 1.48), and hypertension (OR 1.48), all *p* < 0.001. Effect sizes were largest for severe survey-reported HL (1.91) and SNHL (1.36). These findings indicate that severe survey-reported HL and SNHL are strongly associated with dementia.

## Introduction

Dementia encompasses various progressive neurological disorders that impair cognitive function and constitutes a major global health concern, with prevalence estimated to reach 152 million by 2050.^1-3^ The global number of people living with dementia more than doubled from 1990 to 2016 to reach 48 million, highlighting the urgent need for more effective interventions.^4^ Accordingly, there is a growing emphasis on preventative strategies, particularly the early identification and management of potential risk factors, such as hearing loss (HL), cognitive impairment, falls, and depression.^5-10^ Among these, age-related HL has emerged as a major modifiable risk factor, with studies suggesting that its management could reduce the global dementia burden by up to 9%.^11^ Use of hearing aids has been associated with improved hearing function and reduced behavioral symptoms associated with dementia.^11,12^ While most prior studies have focused on audiometrically measured or clinically diagnosed sensorineural HL (SNHL), few have examined both SNHL and self-reported HL simultaneously. SNHL is typically identified via diagnostic codes, whereas survey-reported HL captures participants’ self-perceived hearing difficulties. Our study addresses this gap by evaluating both objectively confirmed and functionally significant hearing deficits as potential early indicators of increased dementia risk, and assessing their value as targets for preventative strategies.

## Materials and Methods

This retrospective case-control study analyzed data from the *All of Us* (AoU) database collected between 1980 and 2022. Approval from an Institutional Review Board was not necessary, given the public nature of the database, informed consent obtained from all survey participants, and the removal of any patient-identifying information from the dataset. The dataset included demographics, dementia diagnoses, comorbidities associated with dementia, and HL, which was captured in two ways. For survey-reported HL, responses were extracted from AoU survey modules that assessed personal and family hearing history, medical diagnoses, and treatment **(Table 1)**. Participants were classified as having severe survey-reported HL if they selected “Self” in response to the survey item: “Including yourself, who in the family has had severe hearing loss or partial deafness in one or both ears?” Because the survey assessed only severe or clinically significant HL, analyses were restricted to this category, and milder or moderate self-reported hearing difficulties were not modeled. For clinically diagnosed SNHL, participants were identified using diagnostic codes for “sensorineural hearing loss” in the electronic health record. Potential dementia cases were identified using diagnostic codes for “impaired cognition,” with dementia diagnosis as the primary outcome. Identified cases were control-matched based on demographic characteristics reflecting the US census. Participants diagnosed with HL after dementia diagnosis were excluded. A total of 16,270 participants were included, with 5,930 diagnosed with SNHL and 1,224 with dementia.

**Table 1.** Original survey-based questions from the *All of Us* database included in analysis.

| **Question** | **Possible Survey Responses** |
| --- | --- |
| How old were you when you were first told you had hearing loss? | Adolescent (12-17), Adult (18-64), Child (0-11), Older adult (65-74)<br>Elderly (75+) |
| Are you seeing a doctor for hearing loss? | Yes, No |
| Including yourself, who in the family has had severe hearing loss or partial deafness in one or both ears? | Daughter, Son, Mother, Father, Grandparent, Sibling, Self |
| Has a doctor or health care provider ever told you that you have any of the following hearing or vision problems? | Cataracts, Nearsighted, Astigmatism, Farsighted, Severe hearing loss or partial deafness in one or both ears, Dry eyes, Glaucoma, Tinnitus, Macular degeneration, Other hearing or eye condition, Blindness, None, Don't Know, Skip Prefer Not To Answer |
| Are you currently prescribed medications and/or receiving treatment for hearing loss? | Yes, No |

A total of 22 variables were included in the regression analysis: 5 demographic variables (gender, age, BMI, race, and Hispanic/Latino ethnicity), 3 dementia-related comorbidities (hypertension, diabetes, and obesity), smoking status, SNHL, HL duration, and 8 survey-based items **(Table 1)**. Data was merged into one dataset using AoU’s Python workbook. The logistic regression model was implemented using Python’s Scikit-learn library. Model performance was assessed using accuracy, sensitivity, specificity, F1 score, and area under the receiver operating characteristic curve (ROC-AUC).

## Results

This study included 16,270 patients, with 6,984 (42.9%) male and 9,029 (55.5%) female participants. Furthermore, 14,283 (87.8%) patients were White, 852 (5.2%) Black or African American, 309 (1.9%) Asian, and 600 (3.7%) Hispanic. The mean age of the cohort was 64.3 ± 14.4 years. A total of 1,224 (7.5%) had a diagnosis of dementia. Severe survey-reported HL was strongly associated with dementia (odds ratio [OR] 6.76, 95% CI 5.04-9.08), followed by SNHL (OR 3.90, 95% CI 3.00-5.06), smoking (OR 1.71, 95% CI 1.20-2.43), having a parent with severe HL (OR 1.48, 95% CI 1.27-1.73), and hypertension (OR 1.48, 95% CI 1.24-1.77), all with p < 0.001 **(Figure 1)**. The ROC-AUC for the predicted occurrence of dementia was 0.711, with an accuracy of 0.722, sensitivity of 63%, and specificity of 96% **(Figure 2)**. Effect sizes (ES) were considered alongside odds ratios to assess the magnitude of associations. Only severe HL by survey (ES 1.91) and SNHL (ES 1.36) exceeded an ES threshold of 1. Given the relatively low prevalence of dementia in the cohort (7.5%), odds ratios were considered a reasonable approximation of relative risk and appropriate for interpreting the strength of association.

**Figure 1.**
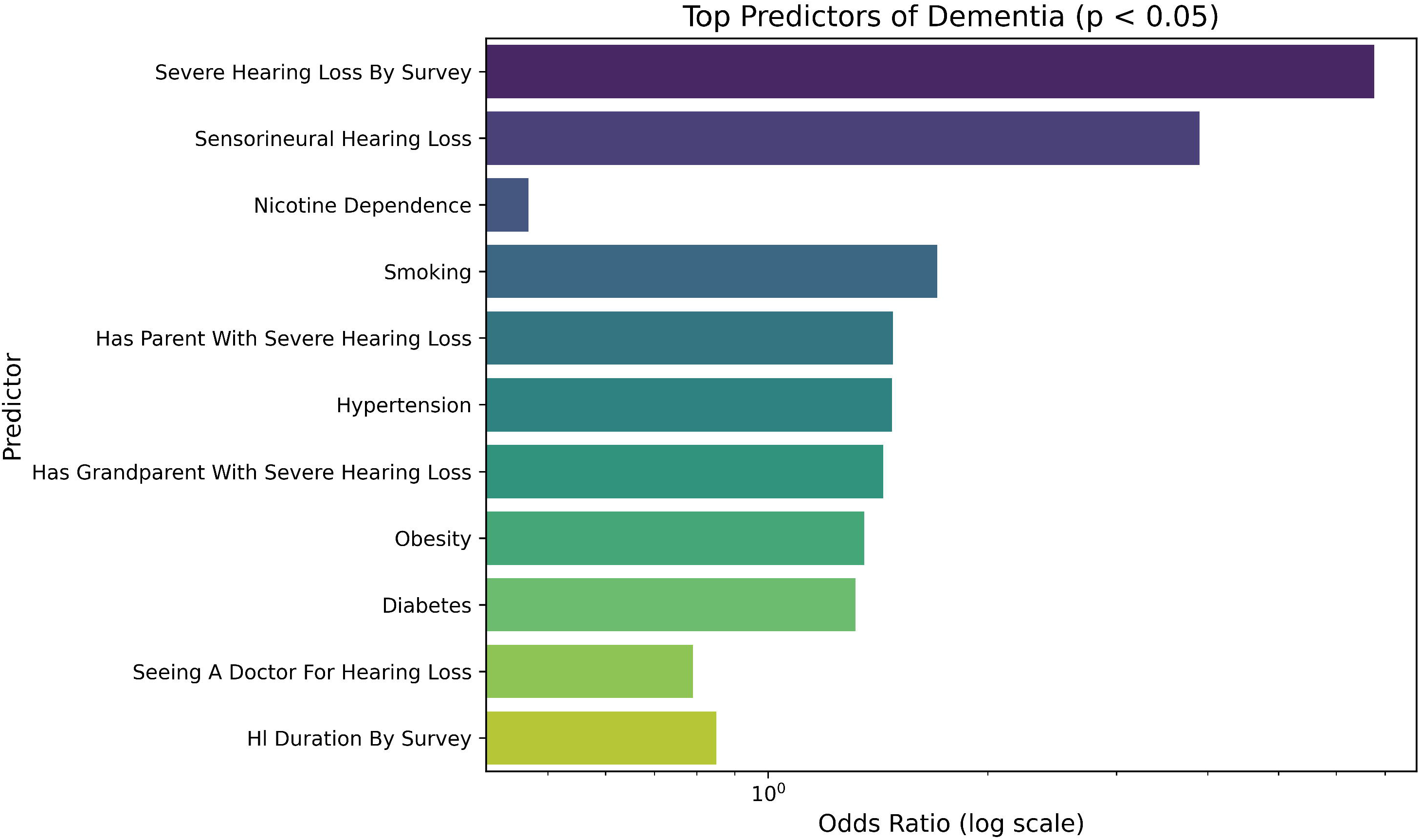
Factors associated with dementia by effect size (Log Odds Ratio)

**Figure 2.**
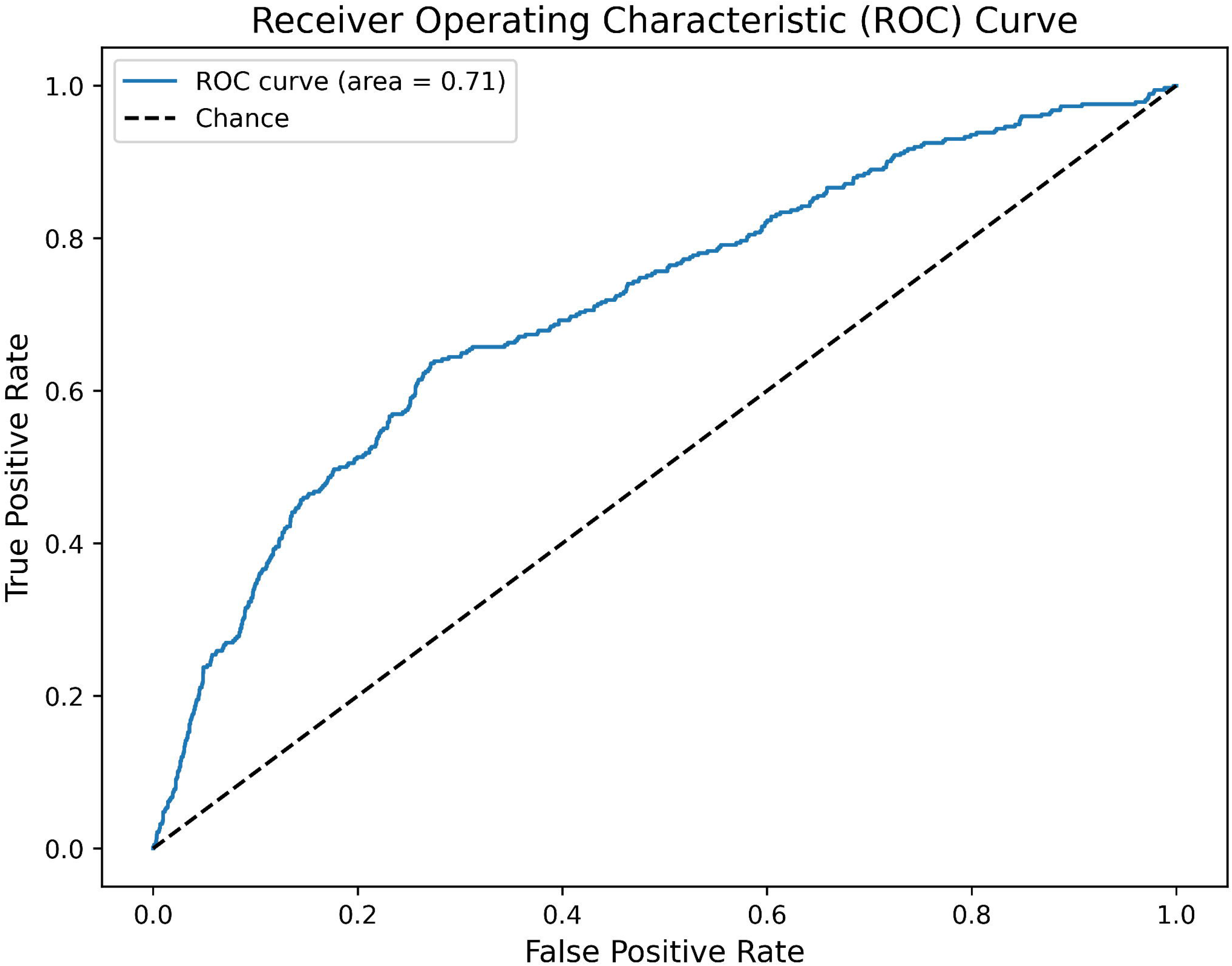
Area under the receiver operating characteristic curve

## Discussion

We found that survey-reported severe HL has the strongest association with dementia, followed by SNHL, smoking, parental HL, and hypertension. Notably, patients reporting severe HL by survey were approximately 7 times more likely to have dementia. Our predictive model demonstrated good discriminatory ability (ROC-AUC 0.711), high specificity (96%), and acceptable accuracy (72.2%). ES analysis further supported these associations, with severe HL and SNHL exceeding the ES threshold of 1, emphasizing their clinical relevance.

These findings align with previous studies suggesting that HL may precede or contribute to cognitive decline.^6-10^ Namely, the social isolation hypothesis suggests that elderly with HL often withdraw from social activities due to communication challenges. Additionally, age-related neural changes may exacerbate the cognitive effects of HL.^3,13^ The strong association with survey-reported HL likely reflects patients’ subjective experience of hearing difficulties, which may better capture the functional impact on daily cognitive challenges than clinical diagnosis alone. A prospective study found that pure-tone audiometry and speech discrimination testing predicted decline in cognitive scores, though the association was not significant enough to predict dementia diagnosis.^14^ In contrast, informant-based HL reported by participants’ study partners were strongly associated with dementia, consistent with prior findings, suggesting central auditory dysfunction beyond peripheral HL.^14^ The relationship between smoking and dementia is well-established in the literature, mediated by increased Alzheimer’s biomarkers, oxidative stress, and neuroinflammation in the cerebrospinal fluid.^15,16^ Hypertension was also found to be a significant modifiable risk factor, reinforcing its known role in cognitive impairment.^17^

Our findings highlight the critical importance of early detection and management of HL as a potential strategy to reduce dementia risk. Interventions including hearing screening, use of amplification devices, smoking cessation, and hypertension control may offer cognitive health benefits, particularly in older adults.^18^ The strength of this study lies in its large sample size and comprehensive dataset, which included both objective and self-reported data. However, there are limitations to be considered when interpreting these findings. The retrospective design limits causal inference because, even after excluding participants diagnosed with HL after dementia, we cannot definitively confirm that HL causes or consistently precedes dementia. Although the AoU Research Program demonstrates improved representation of racial and ethnic minorities compared with historical U.S. biomedical cohorts, our analytic subset underrepresents minority populations, with only 5.2% Black or African American, 1.9% Asian, and 3.7% Hispanic participants. For comparison, the full AoU cohort includes 20.5%, 3.5%, and 16.3% of these groups, respectively, and the corresponding U.S. population proportions are 11.7%, 6.1%, and 16.8%.^19^ This underrepresentation likely reflects multiple factors, including the selection of an analytic subset based on available HL and dementia data, and systemic barriers to minority participation in biomedical research. Prior research in U.S. hearing studies shows similar patterns, with clinical trials reporting predominantly White participants and minimal representation of Black, Asian, and Hispanic/Latino populations.^20,21^ Consequently, our findings may have limited generalizability to minority populations and may not capture potential differences in dementia risk or HL associations across racial and ethnic groups.

## Conclusion

Severe self-reported HL and clinically diagnosed SNHL were linked to dementia, with self-reported HL having the strongest observed association. These findings support the value of early hearing assessment and management, alongside interventions such as smoking cessation and hypertension control, as potential strategies to reduce dementia risk.

## Data Availability

All data produced in the present study are available upon reasonable request to the authors

https://allofus.nih.gov/

## References

1. Booth RG, Dasgupta M, Forchuk C, Shariff SZ. Prevalence of dementia among people experiencing homelessness in Ontario, Canada: a population-based comparative analysis. Lancet Public Health. Apr 2024;9(4):e240–e249.

2. Brookmeyer R, Johnson E, Ziegler-Graham K, Arrighi HM. Forecasting the global burden of Alzheimer’s disease. Alzheimers Dement. Jul 2007;3(3):186–191.

3. Scarinci N, Waite M, Nickbakht M, et al. How do adults with hearing loss, family members, and hearing care professionals respond to the stigma of hearing loss and hearing aids? International Journal of Audiology. 2024;64(Sup1)

4. Collaborators GBDD. Global, regional, and national burden of Alzheimer’s disease and other dementias, 1990-2016: a systematic analysis for the Global Burden of Disease Study 2016. Lancet Neurol. Jan 2019;18(1):88–106.

5. Krellman JW, Mercuri G. Cognitive Interventions for Neurodegenerative Disease. Curr Neurol Neurosci Rep. Sep 2023;23(9):461–468.

6. Golub JS, Brickman AM, Ciarleglio AJ, Schupf N, Luchsinger JA. Association of Subclinical Hearing Loss With Cognitive Performance. JAMA Otolaryngol Head Neck Surg. Jan 1 2020;146(1):57–67.

7. Golub JS, Luchsinger JA, Manly JJ, Stern Y, Mayeux R, Schupf N. Observed Hearing Loss and Incident Dementia in a Multiethnic Cohort. J Am Geriatr Soc. Aug 2017;65(8):1691–1697.

8. Moller S, Lykkegaard J, Hansen RS, Stokholm L, Kjaer NK, Ahrenfeldt LJ. Sensory impairments and the risk of cognitive decline and dementia across sex, age, and regions: Longitudinal insights from Europe. Arch Gerontol Geriatr. Dec 2024;127:105584.

9. Lin FR, Ferrucci L. Hearing Loss and Falls Among Older Adults In the United States. Arch Intern Med. Feb 27 2012;172(4):369–371.

10. Rutherford BR, Brewster K, Golub JS, Kim AH, Roose SP. Sensation and Psychiatry: Linking Age-Related Hearing Loss to Late-Life Depression and Cognitive Decline. Am J Psychiat. Mar 2018;175(3):215–224.

11. Livingston G, Sommerlad A, Orgeta V, et al. Dementia prevention, intervention, and care. Lancet. Dec 16 2017;390(10113):2673–2734.

12. Dawes P, Wolski L, Himmelsbach I, Regan J, Leroi I. Interventions for hearing and vision impairment to improve outcomes for people with dementia: a scoping review. Int Psychogeriatr. Feb 2019;31(2):203–221.

13. Fieldhouse JLP, van Engelen MPE, Handgraaf D, et al. Trajectories of behavior and social cognition in behavioral variant frontotemporal dementia and primary psychiatric disorders: A call for better operationalization of socioemotional changes. European Journal of Neurology. 2024;31(12)

14. Marinelli JP, Lohse CM, Fussell WL, et al. Association between hearing loss and development of dementia using formal behavioural audiometric testing within the Mayo Clinic Study of Aging (MCSA): a prospective population-based study. The Lancet Healthy Longevity. 2022;3(12):e817–e824.

15. Senff J, Tack RWP, Mallick A, et al. Modifiable risk factors for stroke, dementia and late-life depression: a systematic review and DALY-weighted risk factors for a composite outcome. J Neurol Neurosur Ps. Jun 2025;96(6):515–527.

16. Liu YL, Li H, Wang J, et al. Association of Cigarette Smoking With Cerebrospinal Fluid Biomarkers of Neurodegeneration, Neuroinflammation, and Oxidation. Jama Netw Open. Oct 2 2020;3(10)

17. Sierra C. Hypertension and the Risk of Dementia. Front Cardiovasc Med. Jan 31 2020;7

18. Pike JR, Huang AR, Reed NS, et al. Cognitive benefits of hearing intervention vary by risk of cognitive decline: A secondary analysis of the ACHIEVE trial. Alzheimers Dement. May 2025;21(5)

19. Kathiresan N, Cho SMJ, Bhattacharya R, Truong B, Hornsby W, Natarajan P. Representation of Race and Ethnicity in the Contemporary US Health Cohort All of Us Research Program. JAMA Cardiol. 2023 Sep 1;8(9):859–864.

20. Hori K, Jamal M, Zalin M, Li A, Choi JS. Disparities in Clinical Trial Participation for Hearing Loss Treatment and Cognitive Outcomes in the United States: A Scoping Review. The Laryngoscope. 2025 Sep;135(9):3015–3026.

21. Pittman CA, Roura R, Price C, Lin FR, Marrone N, Nieman CL. Racial/Ethnic and Sex Representation in US-Based Clinical Trials of Hearing Loss Management in Adults: A Systematic Review. JAMA Otolaryngol Head Neck Surg. 2021 Jul 1;147(7):656–662.

